# Respiratory syncytial virus (RSV) vaccine effectiveness and antibody correlates of protection among older adults in the Community Vaccine Effectiveness (CoVE) observational study

**DOI:** 10.1101/2025.03.14.25323981

**Authors:** Elie-Tino Godonou, Amy P. Callear, Casey L. Juntila-Raymond, Dolapo Raji, Matthew Smith, Kalee E. Rumfelt, Claire M. Midgley, Leora R. Feldstein, Jefferson M. Jones, Melissa Briggs Hagen, Marisa C. Eisenberg, Adam S. Lauring, Arnold S. Monto, Abram L. Wagner, Emily T. Martin

**Affiliations:** Department of Epidemiology, University of Michigan School of Public Health, Ann Arbor, MI, 48109, United States of America; Coronavirus and Other Respiratory Viruses Division, National Center for Immunization and Respiratory Diseases, Centers for Disease Control and Prevention, Atlanta, GA 30333, USA; Department of Internal Medicine, University of Michigan Medical School, Ann Arbor, MI 48109, United States of America

## Abstract

**Background:** The first RSV vaccines for adults 60 years and older were approved prior to the 2023-2024 respiratory virus season. This study used data from adults 60 years and older, enrolled into the Community Vaccine Effectiveness (CoVE) cohort study, in Michigan, U.S.A, to evaluate RSV vaccine effectiveness (VE) and antibody correlates of protection.

**Methods:** A Cox proportional hazards model was used to compare incidence of symptomatic / all RSV infections in those vaccinated versus unvaccinated. RSV-specific (preF) binding antibodies were measured in serum specimens and assessed longitudinally. A correlates of protection analysis was conducted using logistic regression.

**Findings:** Of the 281 participants (n=117 vaccinated) enrolled (August 1, 2023, to March 1, 2024), 14 tested positive for RSV. Adjusted RSV VE against any RSV infection was 50.8% (95%CI: −79.1% to 86.5%), and 59.8% (95%CI: −105.2% to 92.1%) against symptomatic RSV. There were 61.2 (95%CI: 16.9, 163.2) RSV infections per 1,000 person-years among vaccinated participants compared to 165.8 infections (95%CI: 88.0, 287.0) per 1,000 person-years among those unvaccinated. A 31% decrease in odds (OR: 0.69, 95% CI: 0.44 to 1.07) of RSV infection per 2–fold increase in antibody concentration was observed.

**Interpretation:** RSV infection risk was lower among those with higher antibody titers. RSV incidence was lowest among vaccinated adults, but not significantly. Continued monitoring of reduction of RSV infection in years following vaccination is warranted.

**Funding:** National Center for Immunization and Respiratory Diseases, US Centers for Disease Control and Prevention (75D30122C13149) and National Institute of Allergy and Infectious Diseases (75N93021C00015).

The findings and conclusions in this report are those of the author(s) and do not necessarily represent the official position of the Centers for Disease Control and Prevention.

## Introduction

Considerable evidence has accumulated regarding the significant burden of RSV disease in adults, especially older adults and those with underlying conditions.^1^ A recent U.S.-wide multicenter study of adults hospitalized for acute respiratory illness found that the severity of illness due to RSV was comparable to illness in unvaccinated individuals hospitalized with influenza or COVID.^2,3^ Prevention of RSV infection stands to directly benefit older adults who are at increased risk of severe disease. Simulations suggest that a reduction of infection in this group could lead to indirect reductions in other non-vaccinated groups as well,^4,5^ although this has not been confirmed epidemiologically.

Two protein subunit vaccines for RSV were approved in the U.S. prior to the 2023-2024 respiratory virus season: an adjuvanted vaccine containing recombinant stabilized prefusion RSV-A F protein (AREXVY) and an unadjuvanted vaccine containing recombinant stabilized prefusion RSV-A and RSV-B F proteins combined into a single bivalent product (ABRYSVO). For the first year of implementation, which coincided with our study period, ACIP recommendations for the administration of a single dose of one of the two protein subunit vaccines were for administration to adults 60 and older, based on shared decision making between provider and patient.^6^

Vaccine efficacy estimates from the randomized clinical trials for these vaccines was 62.1% (ABRYSVO) ^7^ and (AREXVY) 71.7%,^8^ for adults 60 and older against RSV-related acute respiratory illness (defined broadly as at least one ^7^ or two ^8^ new or worsening ARI symptom with RT-PCR-confirmed RSV infection). Test-negative evaluations in the U.S. following use in the 2023-2024 season indicated that the real-world vaccine effectiveness against RSV hospitalization in adults 60 and older was 75% (95% CI, 50%-87%).^9^ Broader vaccine effectiveness estimates against less severe outcomes have been limited by the slow uptake of RSV vaccines nationwide.^10^

We sought to evaluate the potential overall benefit of RSV vaccine in older adults during this initial season in a prospective, observational study in a population with high vaccine uptake enrolled across the state of Michigan, The Community Vaccine Evaluation (CoVE) study.^11^ We analyzed RSV symptomatic and asymptomatic infection in vaccinated and unvaccinated cohort participants 60 years of age and older during the 2023-2024 respiratory virus season. Further, we evaluated vaccine immunogenicity, the correlation between antibody levels and RSV infection, and antibody waning in vaccinated and unvaccinated adults.

## Methods

### Study Population and Data Collection

The analysis population included all participating adults in the CoVE cohort who were 60 years of age and over as of their enrollment date, and enrolled at any point during the surveillance period (August 1, 2023, to March 1, 2024). Participant follow-up time was included through the approximate end of RSV circulation in Michigan, March 1, 2024. Individuals were recruited from social media, the University of Michigan research registry, targeted mailing, and an existing household cohort (Household Influenza Vaccine Evaluation Study [HIVE], methods previously described^12,13^). In anticipation of RSV vaccine availability, targeted social media recruitment was used beginning 6/20/2023 to increase enrollment of participants aged 60 and above throughout the state. Adults provided written informed consent. This study was reviewed and approved by the University of Michigan Institutional Review Board, and was conducted consistent with applicable federal law and CDC policy (See 45 C.F.R. part 46.114; 21 C.F.R. part 56.114).

Participant and household characteristics, health history, and vaccination history were collected at enrollment; self-reported data were used to document the presence of high-risk health conditions (i.e., any of these conditions: asthma, chronic obstructive pulmonary disease including chronic bronchitis and emphysema, heart disease, diabetes, chronic kidney disease or loss of kidney function, chronic liver disease such as hepatitis, a bleeding disorder or a blood disease such as sickle cell anemia, weakened immune system, and a neurologic or neuromuscular disorder such as cerebral palsy, multiple sclerosis, or myasthenia gravis). RSV vaccination, including date of vaccination and product, was documented using the Michigan Care Improvement Registry (a statewide immunization information system),^14^ at the conclusion of the study period.

Participants self-collected mid-turbinate swabs weekly, returned them to the study site by mail, and completed weekly surveys on illnesses and individual symptoms (cough, chills, fever, sore throat, nasal congestion, body aches, headache, trouble breathing, wheezing, fatigue). An additional swab was collected upon onset of eligible symptoms meeting the study illness case definition (two or more of: fever, chills, cough, nasal congestion, body aches, headache, sore throat) with symptoms recorded at illness report. A blood sample was requested upon enrollment, with optional additional collections semi-annually and following COVID-19 vaccination. Samples were secondarily used to study RSV vaccination if timed appropriately (see inclusion criteria below). Blood samples were self-collected using an at-home capillary collection kit (TASSO+ SST, Tasso, Inc. Seattle, WA) or collected by study staff or clinical phlebotomist via venous draw. The median time from collection to receipt in the laboratory was 2 days for venous samples (n=112; centrifuged day of collection) and 3 days for home-collected samples (n=400). Home-collected samples were returned via mail; samples were discarded if serum failed to appropriately separate after centrifugation. Hemolyzed serum was not excluded from analysis.

### Laboratory Testing

Dry nasal swabs received from participants were transferred into 2ml stabilizing solution (DNA/RNA Shield, Zymo Research Corporation, Irvine, CA) upon receipt in the study laboratory. All respiratory specimens were tested by real-time reverse-transcription polymerase chain reaction (RT-PCR) for detection of RSV virus using a multiplex assay for RSV, SARS-CoV-2, and influenza (Taqman SARS-CoV-2, Flu A/B, RSV Multiplex Assay, AppliedBiosystems, Waltham, MA) as previously described.^11^

Available serum specimens were tested for IgG antibody against the RSV Pre-Fusion F protein using a quantitative electrochemoluminescence assay (V-PLEX Respiratory Panel 1 (IgG), Mesoscale Discovery, Rockville, MD). Specimens were tested in duplicate using a 1:5,000 dilution with retesting at 1:50,000 if the first test was above the upper limit of quantitation. Absolute antibody quantity was determined using a standard curve, in arbitrary units per mL (AU/mL).

### RSV antibody kinetics

The kinetics of RSV preF IgG levels in the vaccinated and unvaccinated groups were characterized using multiple models, based on blood specimens collected between August 1, 2023, to July 15, 2024.

Among vaccinated participants, individuals with at least two post-vaccination samples collected at least 14 days after RSV vaccination were selected. If any vaccinated participants became infected (PCR-confirmed RSV, regardless of symptoms) after vaccination, antibody levels measured after infection were excluded from the waning analysis.^15^

Among unvaccinated participants, individuals with at least two samples collected at different time points after August 1, 2023, were selected. The same method described above (i.e., with exclusion of post-infection samples) was applied if any unvaccinated participants were infected (PCR-confirmed RSV, regardless of symptoms). Post-infection antibody kinetics were not assessed due to fewer PCR-confirmed RSV cases with associated samples.

### Statistical Analysis

A descriptive analysis was performed to characterize participants who received RSV vaccination during the study period and those who did not, including demographic characteristics, RSV infections (any positive swab, inclusive of symptomatic *and* asymptomatic detections), and symptomatic RSV illness (a positive test and illness symptoms meeting the case definition above; henceforth referred to as “symptomatic RSV”). Symptom frequency and intensity were described among participants with symptomatic RSV. Symptomatic RSV infection was defined as a positive RT-PCR test and the presence of at least two respiratory symptoms reported within seven days before or after the specimen collection date. Repeated RSV positives observed in the same participant within 14 days were considered a continuation of the prior RSV infection. Asymptomatic cases were defined as those that did not meet the symptomatic RSV case definition, and may have included individuals with other non-qualifying symptom combinations. Participants contributed time under observation to the vaccinated group if they were vaccinated at least 14 days prior to either any RSV infection, or the end of the study period.

A Cox Proportional-Hazards (CPH) model, with a robust sandwich estimate for the covariance matrix, was used to evaluate vaccine effectiveness in reducing the hazard rate of RSV infections (symptomatic infections only and all cases of RSV infection) over time, adjusted for sex, race, ethnicity, and presence of high-risk health conditions (a binary yes/no variable).^2,11,16–19^ Vaccination status was a time-varying covariate and time to infection was measured from August 1, 2023, or participant enrollment date if later, with the Efron approximation method used for ties in event times. Vaccine effectiveness (VE) was estimated as (1 − Hazard ratio) × 100.

Unadjusted incidence rates (IR) per 1,000 person-years, with 95% confidence intervals (CIs),^20^ were calculated separately for RSV infection and symptomatic RSV overall and stratified by vaccination status. Person-years were calculated up to the occurrence of the first event, loss to follow up, or the end of the study period.

We also fit an unadjusted logistic regression model to evaluate continuous levels of RSV antibody on the log scale as a correlate of protection (CoP) in our study population. This model estimated the change in the odds of RT-PCR RSV infection associated with each 2-fold rise in baseline titers. This CoP analysis included vaccinated and unvaccinated participants combined (Supplemental Materials).

For the immunogenicity assessment, Geometric Mean Fold Rise (GMFR) with a 95% confidence interval (CI) was calculated for participants with both pre- and post-vaccination titer results.

We modeled RSV preF IgG waning from 14 - 302 days after vaccination among vaccinated individuals, and from 1 - 349 days after the start of the study period among unvaccinated, based on the timeframe of available specimens within one year of vaccination. RSV preF IgG waning predicted trajectories were estimated using exponential decay (ED), power-law (PL), and natural cubic splines (NCS) mixed models, starting from day 14 post-vaccination for the vaccinated group and from day 1 after the start of the study period for the unvaccinated group. All waning models were implemented using the lme4 package in R,^21^ and 95% confidence intervals of the fixed effects from ED and PL mixed models are calculated by computing a likelihood profile and finding the appropriate cutoffs based on the likelihood ratio test.^21^ A description of these three models is provided in the supplemental materials.

Antibody half-life (t_1/2_), representing the number of days required to halve RSV preF IgG antibody levels, was estimated for both vaccinated and unvaccinated groups for each model (Supplemental Materials). The confidence intervals for the estimated half-lives were computed using a nonparametric bootstrap method as implemented in the boot package in R.^22,23^ All analyses were conducted using SAS software (version 9.4; SAS Institute, Cary, NC) and R software (version 4.3.0; R Core Team).

## Results

A total of 281 participants aged 60 years and older participated in the study between August 1, 2023, to March 1, 2024, and contributed 5733 respiratory specimens (weekly and illness swabs) and 115.38 person-years of person-time to the analyses (Table 1). Overall, participants were predominantly female (71.2%, n=200) and White (93.6%, n=263) with a median age of 67 years (range, 60 - 87); a large proportion (90.7%, n=255) of participants belonged to the age category 60-74 years. One third of participants (33.1%, n=93) had at least one high-risk health condition, and 5% (n=14) had a PCR-confirmed RSV infection (Table 1).

**Table 1.**
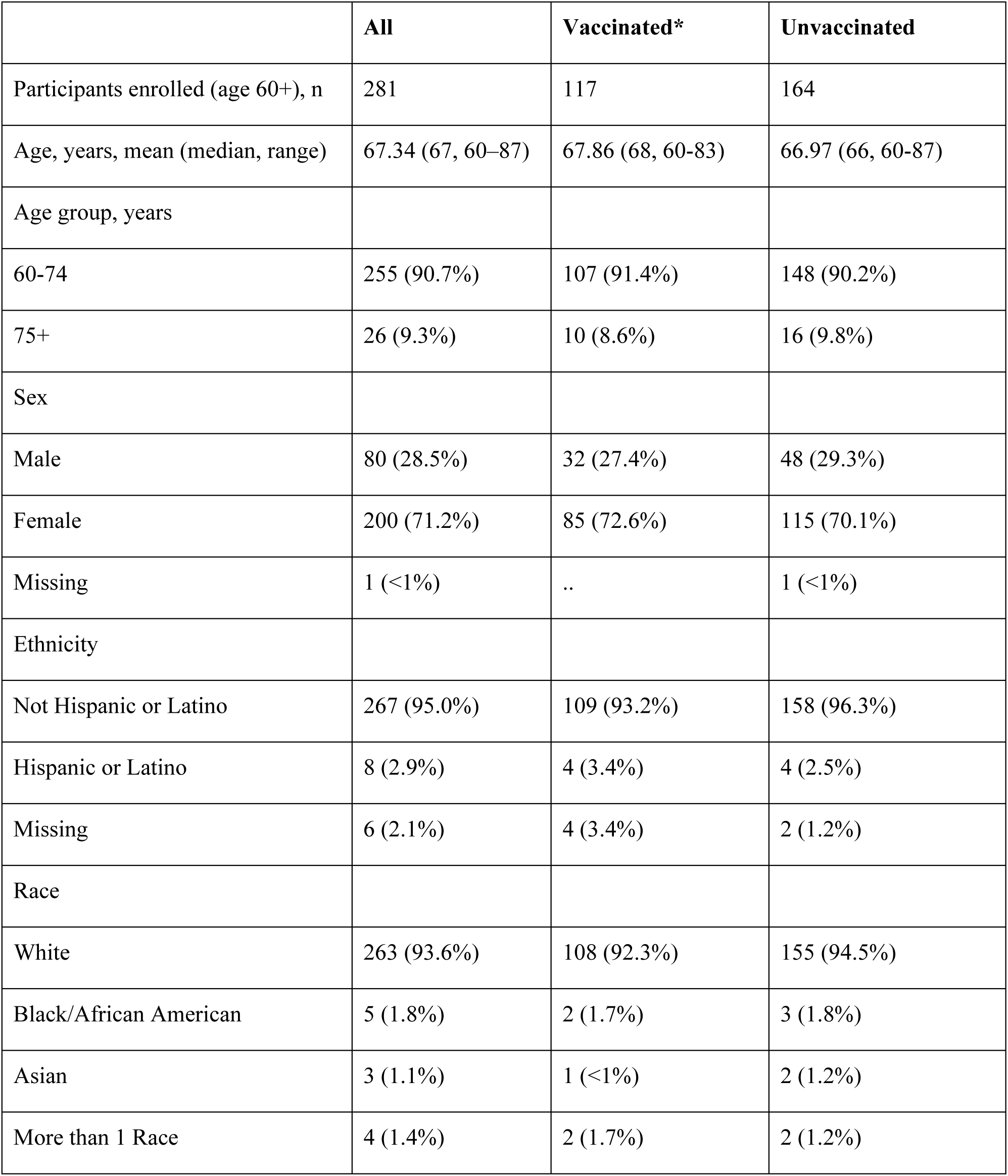
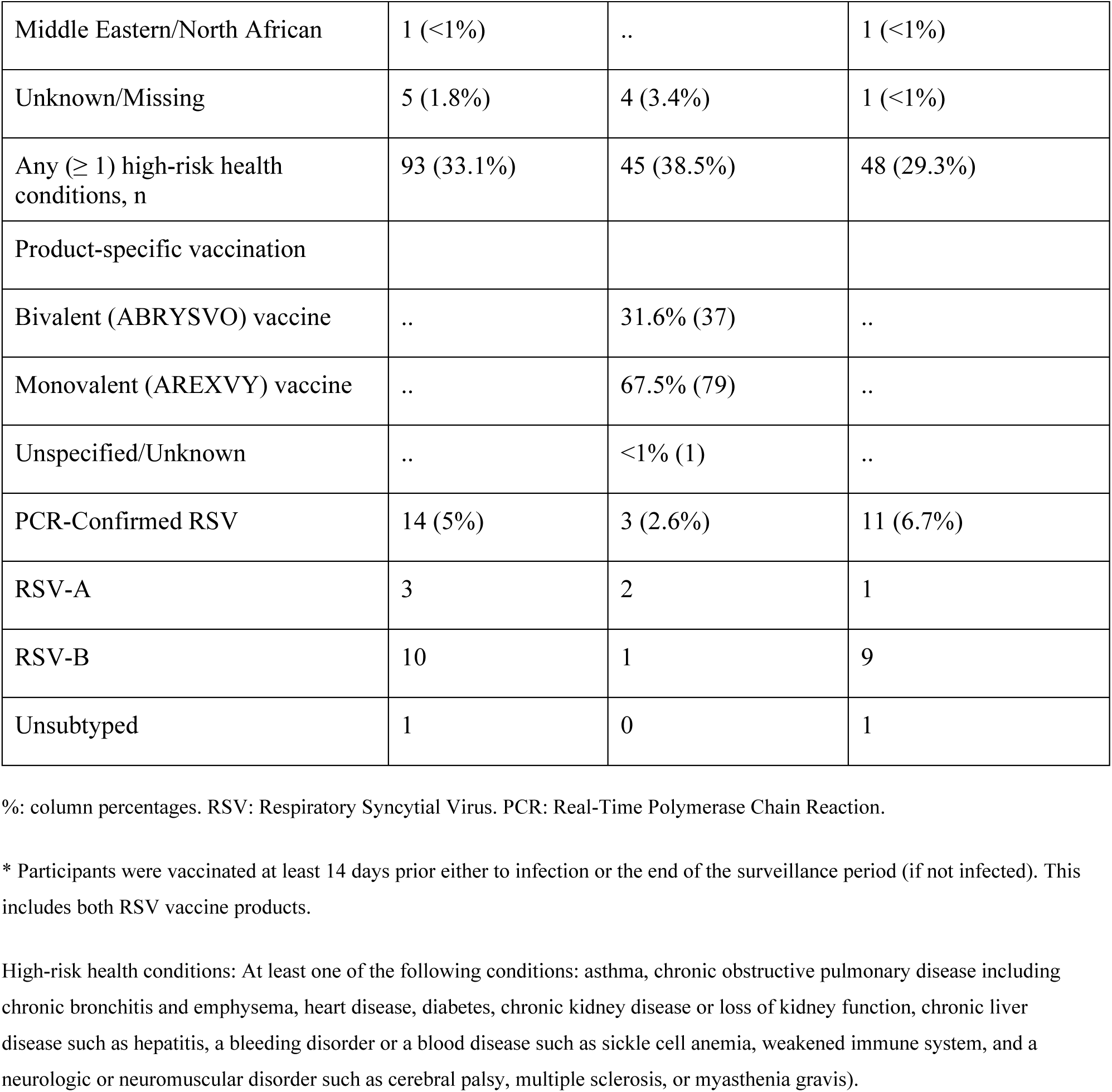
Characteristics (Age, Sex, Race, High-Risk Health Conditions) of Participants Aged 60 and Older, PCR-Confirmed RSV Infections, and RSV Subtypes, Grouped by RSV Vaccination Status.

Among the 14 RSV-infected participants, ten had the RSV-B antigenic subtype, three had the RSV-A subtype, and one sample was unsubtypeable. Among symptomatic RSV cases (3.6%, n=10), all participants experienced cough and nasal congestion, while nine also reported a sore throat (Table 2); only one (unvaccinated) participant missed work, due to the illness, for two days. No symptomatic RSV cases required hospitalization or visits to an emergency department or urgent care. Only one participant had an outpatient healthcare visit (Table 2). Of the 14 participants with RSV infection, 3 (21.4%) were vaccinated; of these three individuals, two were symptomatic, and none sought medical care. In the two vaccinated cases with symptomatic infection, illness onset occurred 80 and 145 days following vaccination. In the third individual, asymptomatic RSV detection occurred by RT-PCR in a weekly swab 68 days following vaccination.

**Table 2.** Frequency of Symptoms, Outpatient Healthcare Visits, Emergency Department or Urgent Care Visits, and Hospitalizations in Participants Aged 60 and Older with Symptomatic RSV Infection Grouped by Vaccination Status.

|  | **All** | **Vaccinated\*** | **Unvaccinated** |
| --- | --- | --- | --- |
| Symptomatic RSV, n | 10 | 2 | 8 |
| Symptoms |  |  |  |
| Cough | 10 (100%) | 2 (100%) | 8 (100%) |
| Chills | 4 (40%) | 0 | 4 (50%) |
| Fever | 3 (30%) | 0 | 3 (37.5%) |
| Sore Throat | 9 (90%) | 1 (50%) | 8 (100%) |
| Nasal Congestion | 10 (100%) | 2 (100%) | 8 (100%) |
| Body Aches | 5 (50%) | 0 | 5 (62.5%) |
| Headache | 8 (80%) | 2 (100%) | 6 (75%) |
| Trouble Breathing | 1 (10%) | 1 (50%) | 0 |
| Wheezing | 4 (40%) | 1 (50%) | 3 (37.5%) |
| Fatigue | 8 (80%) | 1 (50%) | 7 (87.5%) |
| Outpatient Healthcare Visit | 1 | 0 | 1 |
| Emergency Department or Urgent Care Visit | 0 | 0 | 0 |
| Hospitalization (overnight) | 0 | 0 | 0 |

%; column percentages.
\* Participants were vaccinated at least 14 days prior either to infection or the end of the surveillance period (if not infected). This includes both RSV vaccine products.

A total of 41.6% (n=117) of included participants received an RSV vaccine, and 91.4% (n=107) of those vaccinated were in the 60-74 age category. Importantly, vaccine uptake coincided with circulation of RSV in the broader cohort (Figure 1). Regarding product-specific vaccination, 67.5% received GlaxoSmithKline’s monovalent (AREXVY) vaccine, while 31.6% received the Pfizer’s bivalent (ABRYSVO) vaccine among those vaccinated (Table 1).

**Figure 1.**
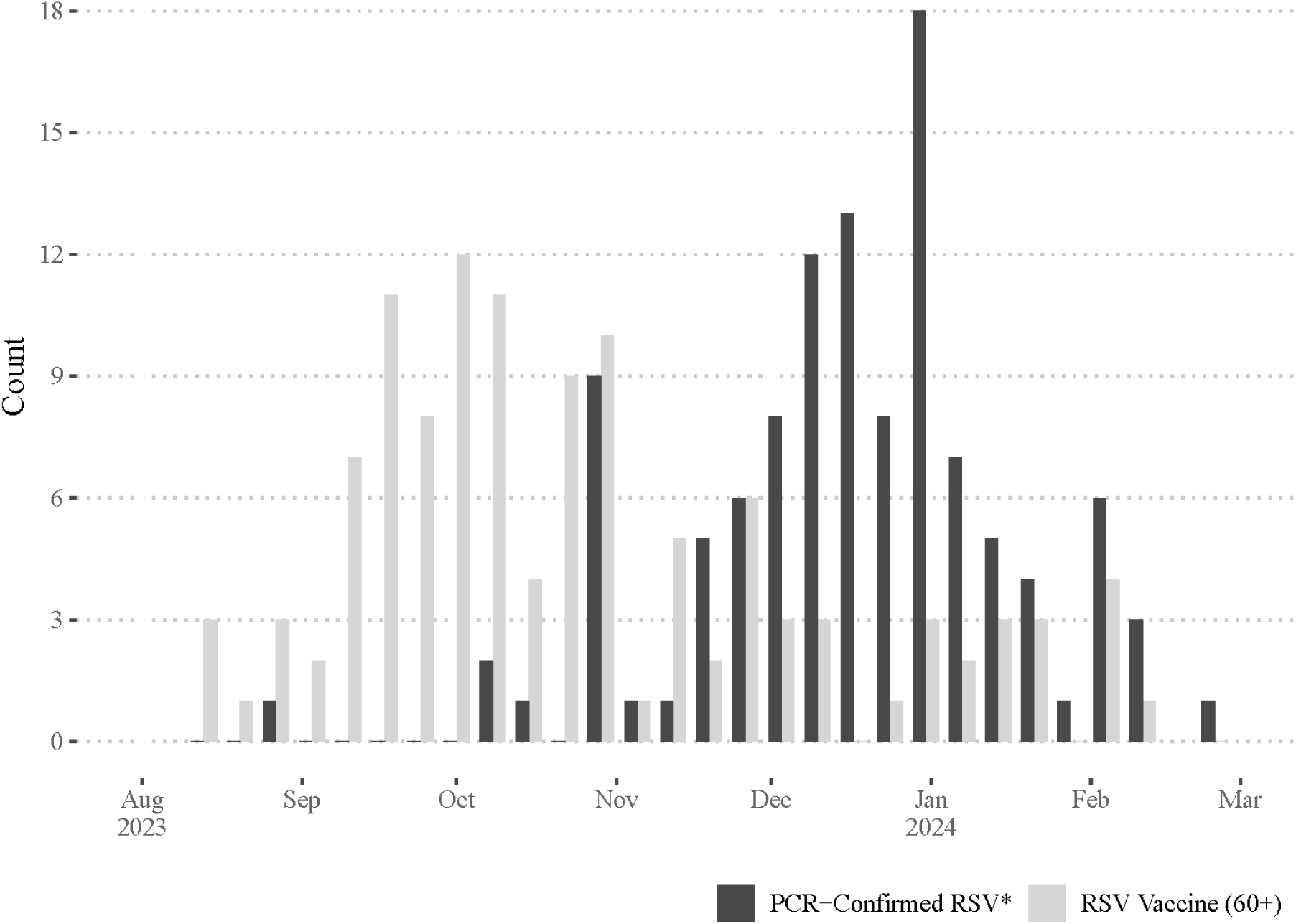
COVE RSV vaccinations (light gray) in adults 60 years of age and older, included in the analysis. All RT-PCR-confirmed RSV infections (dark gray) in all CoVE participants of any age during the 2023-2024 season are noted in green. *RSV among 994 participants enrolled in COVE. 112 RSV infections among 108 people (4 people with 2 infections). Median age 13.6 years (1 - 83). RSV: Respiratory Syncytial Virus. RT-PCR: Real-Time Polymerase Chain Reaction.

The unadjusted incidence rate (IR) of all RSV infections, inclusive of symptomatic and asymptomatic cases, was 121.3 (95%CI: 69.5, 198.1) per 1,000 person-years. For RSV cases that did not meet the symptomatic case definition, IR was 34.7 (95%CI: 11.6, 82.4) per 1,000 person-years, which was lower than that for symptomatic RSV cases (86.7; 95%CI: 44.5, 153.7); this difference was not statistically significant. IR among unvaccinated participants was higher than vaccinated participants, however the difference was not statistically significant. Overall, there were 61.2 (95%CI: 16.9, 163.2) RSV infections per 1,000 person-years among vaccinated participants compared to 165.8 infections (95%CI: 88.0, 287.0) per 1,000 person-years among those unvaccinated (Table 3); the incidence of symptomatic RSV among unvaccinated was 120.6 infections (95%CI: 56.9, 227.6) per 1,000 person-years compared to 40.8 infections (95%CI: 8.1, 130.7) per 1,000 person-years among vaccinated participants. The adjusted RSV VE against any RSV infection was 50.8% (95%CI: −79.1% to 86.5%) and 59.8% (95%CI: −105.2% to 92.1%) against symptomatic RSV (Table 3).

**Table 3.**
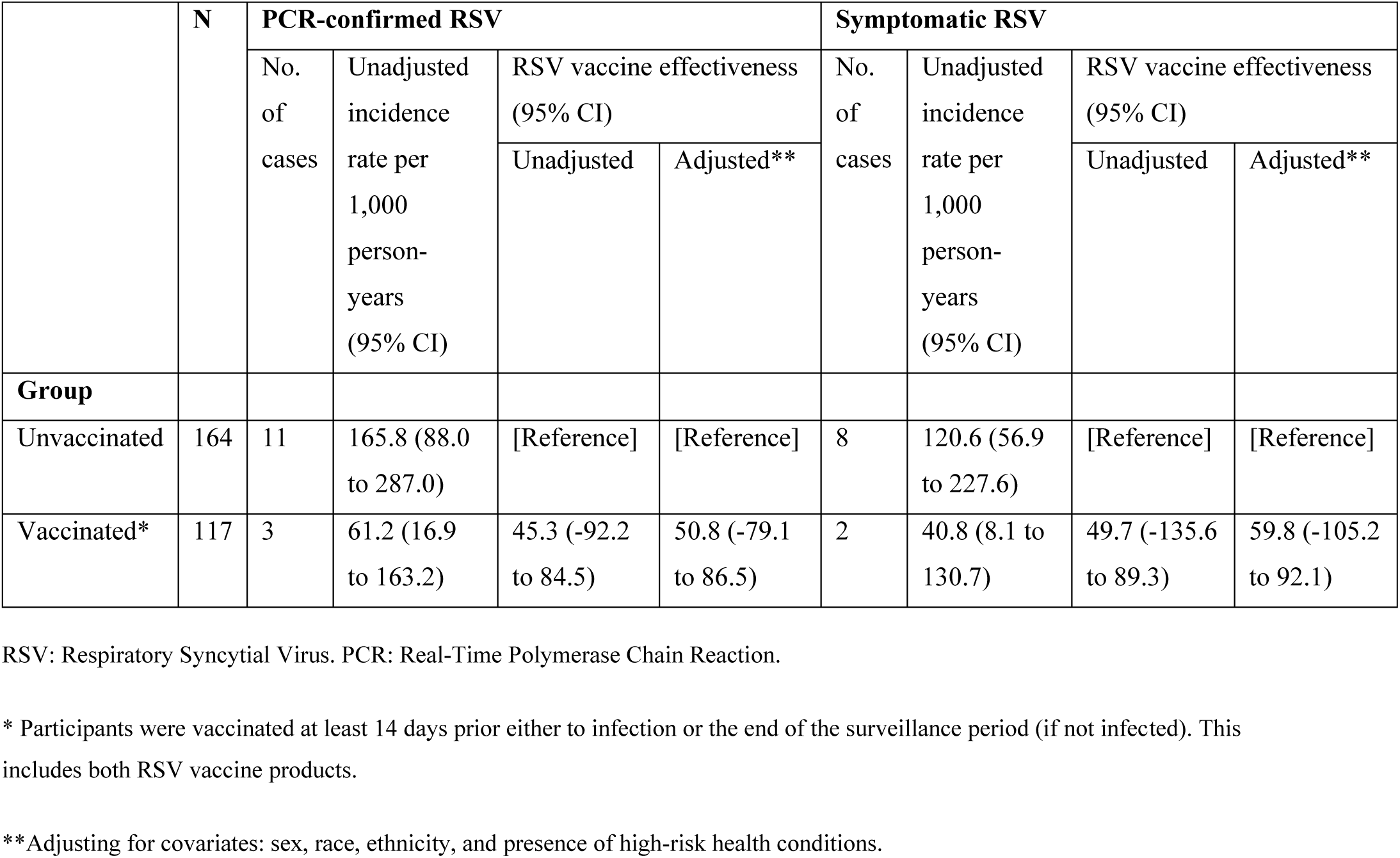
RSV Vaccine Effectiveness Against PCR-Confirmed or Symptomatic RSV Infection Among Older Adults.

Geometric mean fold rise (GMFR) for antibody titers against RSV was 14.03 (95%CI: 10.1 - 19.5) among 35 participants who had blood specimens collected before and after their vaccination; 32 participants had at least a 4-fold rise (range, 4.0 - 126.8) in antibody titer. Overall, risk of RSV infection decreased as the titer of IgG antibody against RSV pre-fusion F increased (Figure 2), this analysis included both vaccinated and unvaccinated participants. In total, 13 infected and 81 uninfected participants were included in the CoP analysis, after baseline sample selection. A 31% decrease in odds (Unadjusted OR: 0.69, 95% CI: 0.44 to 1.07) of RSV infection per 2-fold increase in baseline titers was observed but was not statistically significant. A 53% decrease in odds, not significant, was observed (unadjusted OR: 0.47, 95% CI: 0.19 to 1.15) per 4-fold increase in baseline titers.

**Figure 2.**
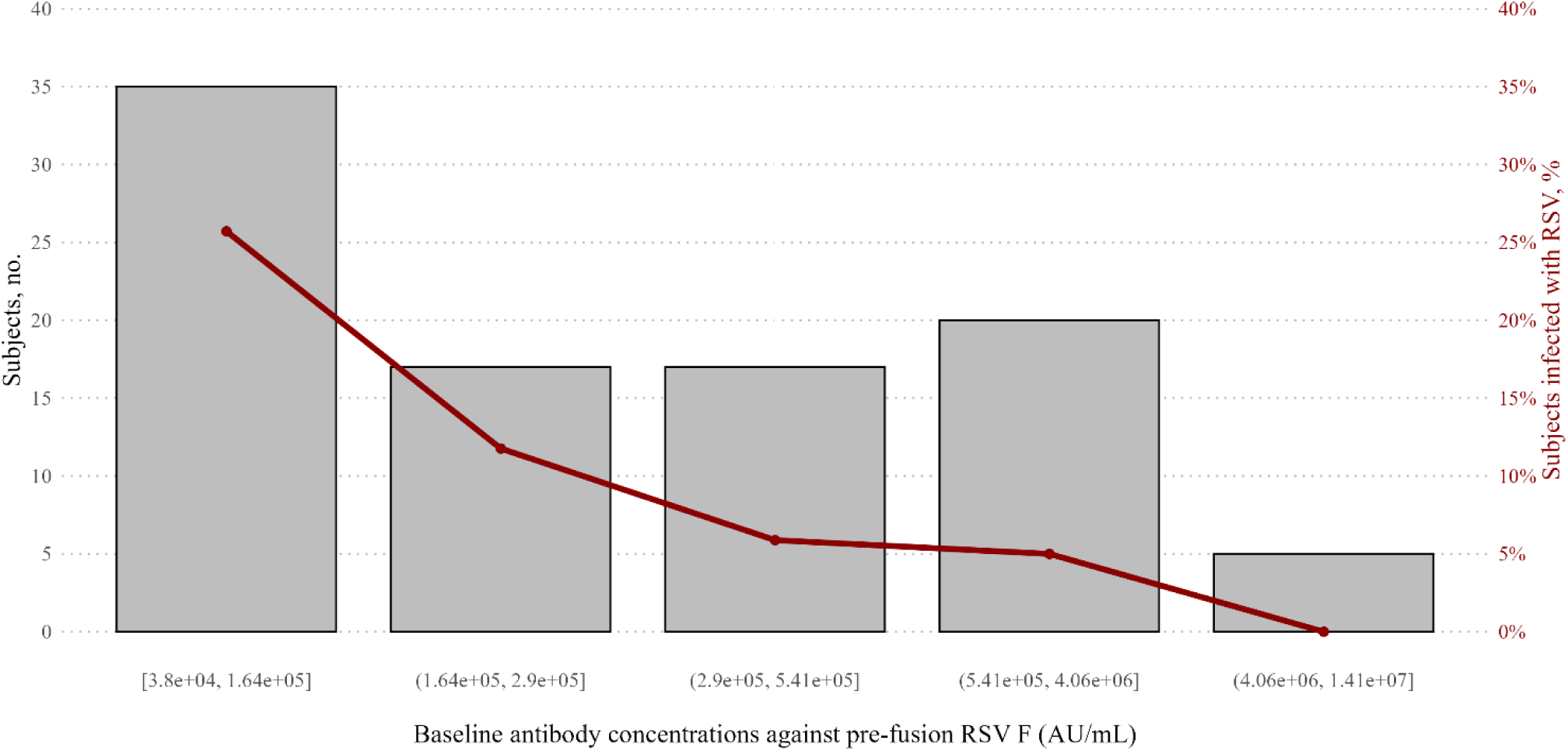
Relationship between measured antibody concentrations against pre-fusion RSV F protein and RSV infection risk. The number of subjects within each interval is plotted on the left y-axis, and the proportion of subjects who were infected with RSV is plotted on the right y-axis, with the red line denoting the proportion infected. RSV: Respiratory Syncytial Virus. AU/ml: Arbitrary Units per ml.

Overall, 54 individuals were included from the vaccinated group and 95 from the unvaccinated group, for the analysis of longitudinal changes in RSV preF IgG levels on a log_10_ scale over time since vaccination among the vaccinated (14 days+) and time since the start of the study period for the unvaccinated (Figure 3). The decay rates (with 95%CI), AICc and BIC criteria from ED and PL mixed models are reported in Table 4. NCS mixed models’ AICc criteria for the vaccinated and unvaccinated groups were 80.3 (with BIC = 96.5) and 337.5 (with BIC = 357.7) respectively (Table 4). Based on these criteria, the best fit for the vaccinated group was the NCS mixed model, with an AICc value of 80.3, compared to 126 and 139.2 for the ED and PL mixed models, respectively. Likewise, the NCS mixed model was the best fit for the unvaccinated group (Table 4). We explored the pattern of RSV preF IgG waning over time using the three models described above. We compared the predicted trajectories of RSV preF IgG levels on a log_10_ scale over time based on the three models above (Figure 4). According to the NCS mixed model, the number of days required to halve RSV preF IgG antibody levels, compared to the levels on the 14th day after vaccination, was 37.8 days (95%CI: 23.3, 145.5). In the ED (i.e., steady decay rate over time) and PL (i.e., decay rates decrease over time) mixed models, this was 72.7 days (95%CI: 61.1, 90.2), and 17.5 days (95%CI: 14.5, 26.2), respectively (Table 4).

**Figure 3.**
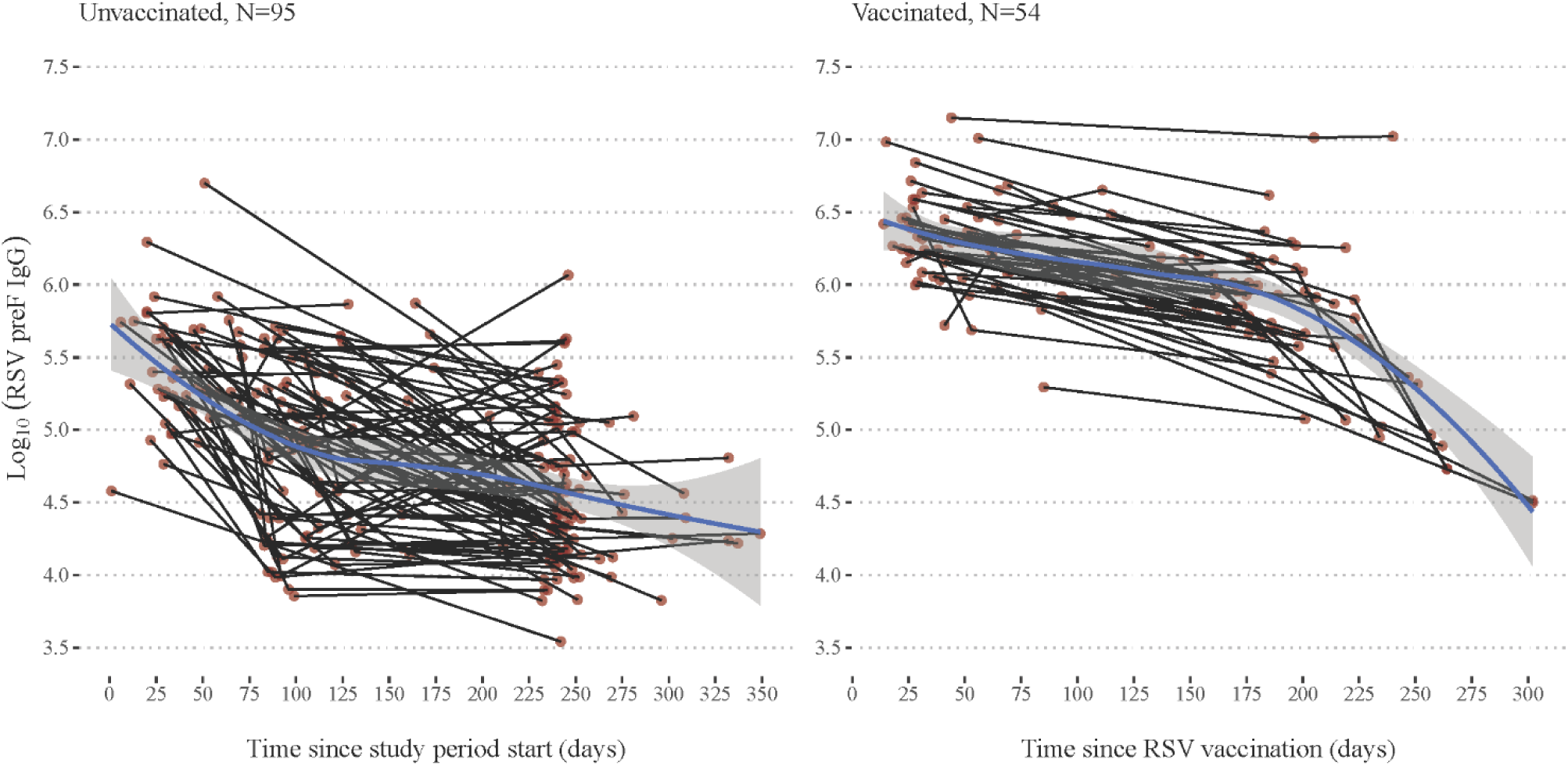
Spaghetti plots of longitudinal changes in RSV preF IgG concentrations over time. The time scale on the left panel (unvaccinated participants) corresponds to the time since the start of the study period (derived from the timing between August 1, 2023, and sample collection date). The time scale on the right panel (vaccinated participants) is time since RSV vaccination (derived from the timing between RSV vaccination date and sample collection date). A loess curve was added to each panel to examine the overall trend. N= number of individuals. RSV: Respiratory Syncytial Virus.

**Figure 4.**
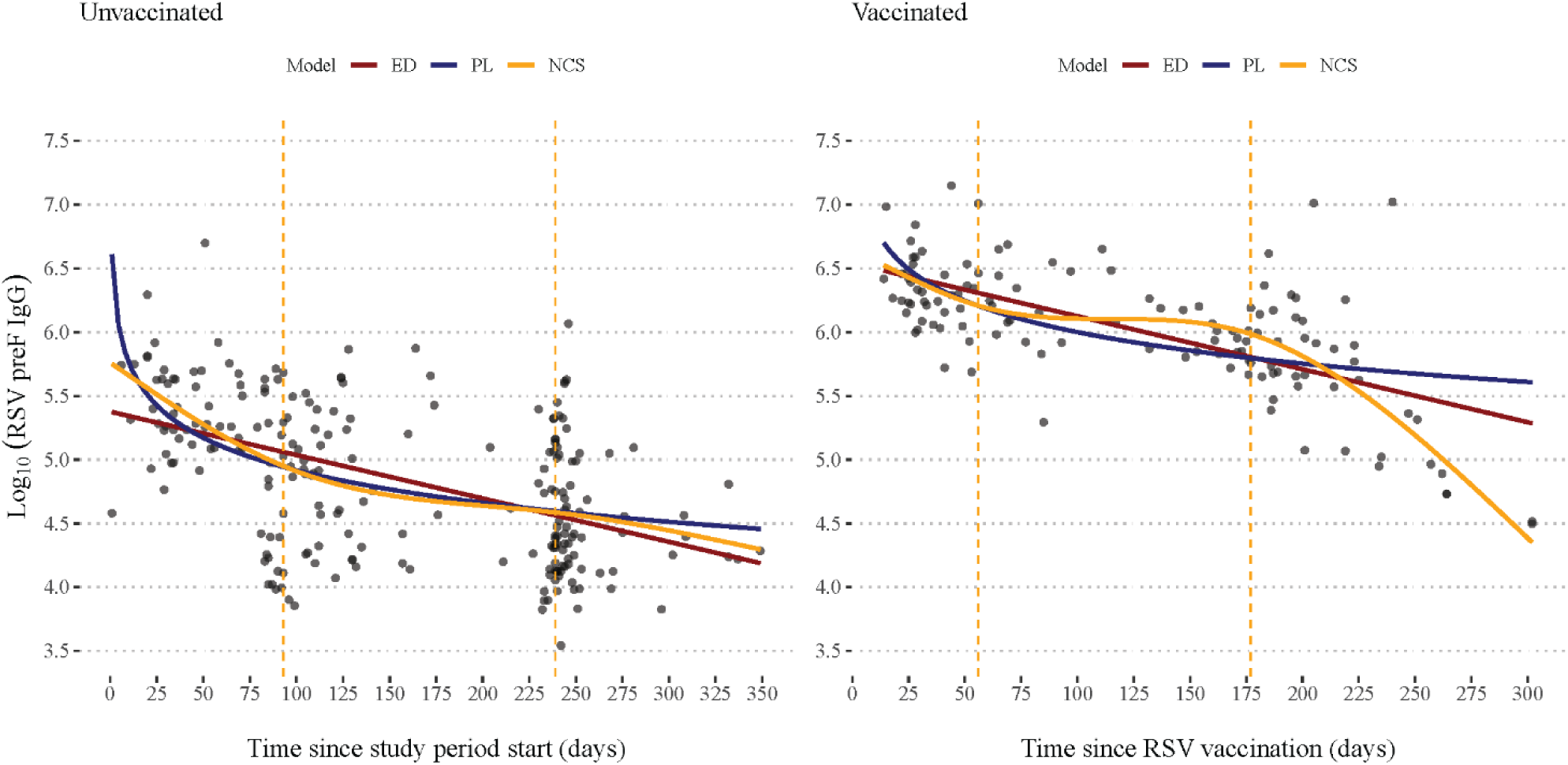
Predicted trajectories of RSV preF IgG levels over time using ED (Exponential Decay), PL (Power-Law), and NCS (Natural Cubic Splines) mixed models. Dots on both panels represent the original data points on a log10 scale. The vertical dashed lines represent the location of the two inner knots from the NCS mixed models. RSV: Respiratory Syncytial Virus.

**Table 4:**
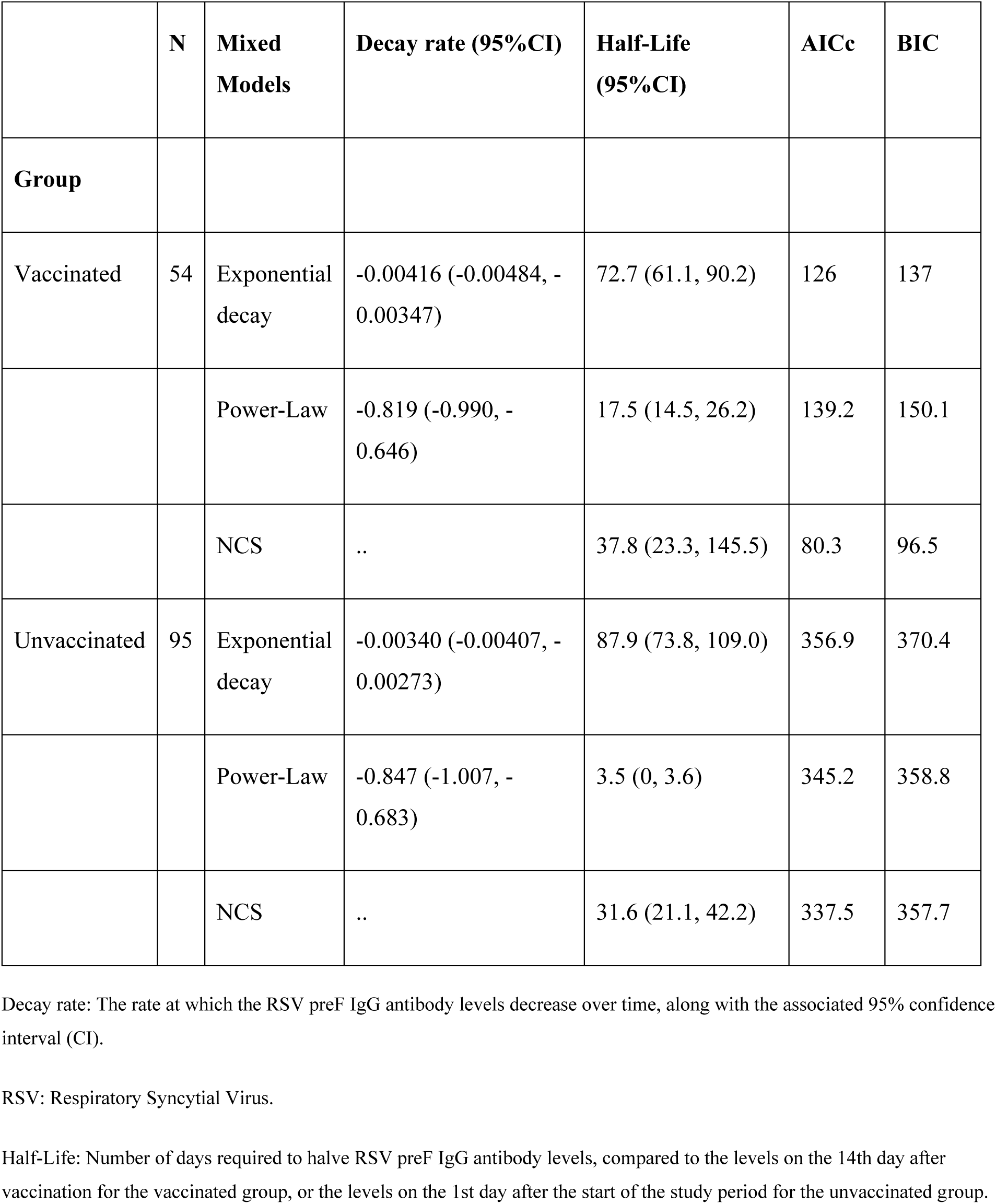
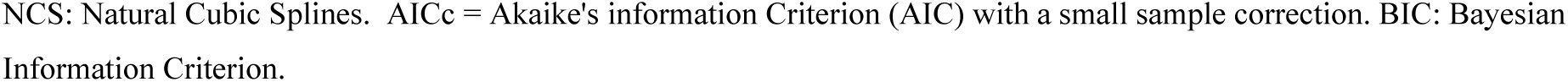
Time (Days) Required to Halve RSV preF IgG Antibody Levels: Estimates from Exponential Decay, Power-Law, and Natural Cubic Splines Mixed Models.

|  | N | Mixed Models | Decay rate (95%CI) | Half-Life (95%CI) | AICc | BIC |
| --- | --- | --- | --- | --- | --- | --- |
| Group |  |  |  |  |  |  |
| Vaccinated | 54 | Exponential decay | -0.00416 (-0.00484, -0.00347) | 72.7 (61.1, 90.2) | 126 | 137 |
|  |  | Power-Law | -0.819 (-0.990, -0.646) | 17.5 (14.5, 26.2) | 139.2 | 150.1 |
|  |  | NCS | .. | 37.8 (23.3, 145.5) | 80.3 | 96.5 |
| Unvaccinated | 95 | Exponential decay | -0.00340 (-0.00407, -0.00273) | 87.9 (73.8, 109.0) | 356.9 | 370.4 |
|  |  | Power-Law | -0.847 (-1.007, -0.683) | 3.5 (0, 3.6) | 345.2 | 358.8 |
|  |  | NCS | .. | 31.6 (21.1, 42.2) | 337.5 | 357.7 |

Decay rate: The rate at which the RSV preF IgG antibody levels decrease over time, along with the associated 95% confidence interval (CI).
RSV: Respiratory Syncytial Virus.
Half-Life: Number of days required to halve RSV preF IgG antibody levels, compared to the levels on the 14th day after vaccination for the vaccinated group, or the levels on the 1st day after the start of the study period for the unvaccinated group.
NCS: Natural Cubic Splines. AICc = Akaike's information Criterion (AIC) with a small sample correction. BIC: Bayesian Information Criterion.

## Discussion

We present an early look at RSV infection and symptomatic illness among vaccine-eligible, community-dwelling adults during the 2023-2024 respiratory season. Vaccine uptake in the cohort was high (42%) compared to 17% reported nationally,^10^ serologic antibody responses to vaccine administration were uniformly strong, and RSV infection risk was lower among those with higher antibody titers. Incidence of RSV infection was lowest among vaccinated groups, though not significantly different. Three vaccinated individuals had subsequent RSV infection in our study. Little is currently known about mechanisms that might lead to RSV infection following vaccination, and prospective studies such as this have valuable baseline samples and participant information that can shed light on this question.

The RSV illnesses observed in this study were mild, rarely requiring medical care, and sometimes asymptomatic. The importance of asymptomatic infection in RSV transmission is unknown. To that effect, the degree to which RSV vaccination of older adults may confer indirect protection to other vulnerable individuals is an important consideration for public health. Overall, the mild nature of illnesses in this study was unsurprising given the weekly participation that may be too intensive for someone with a high level of multimorbidity and functional limitations, both risk factors for more severe RSV illness.^24^

In June 2024, the ACIP updated their recommendation for RSV vaccine use in older adults.^25^ The revised guidance recommended RSV vaccine for all adults 75 years of age or older, and adults 60-74 with risk factors for severe disease. The age distribution of our study population is on the younger end of the age range from the original 2023 guidance^6^ and, on average, younger than recent epidemiologic analyses of severe RSV illnesses.^9^ The importance of understanding RSV epidemiology, and potential prevention strategies in this age group, is twofold. First, the vaccine recommendation for those 60 to 74 years of age is for those at high risk of severe RSV disease. Our group and others have described over many years the potential burden of disease and severe outcomes of RSV that can occur for adults with chronic conditions even if they are in their 50’s and 60’s. One-third of our study population reported at least one high-risk health condition. Second, adults 60 to 74 are frequently still in the workforce and as well as providing direct care for their children or grandchildren. Other studies have shown that RSV can lead to missed days of work in over a third of infections in adults,^26^ and the economic impact of mild illnesses can still be substantial even when medical care is not required.

The CDC currently only recommends one dose of RSV vaccine for eligible adults, without a recommendation for repeating doses in subsequent RSV seasons.^6^ It is too early to know whether this durability over multiple years will hold true in real-world settings outside the context of a clinical trial, highlighting a need for continued assessment. In addition, future efforts will need to account for the fact that additional products have been approved prior to the 2024-2025 season. Thus, continued monitoring of real-world RSV vaccine effectiveness and immunologic waning in the coming years will be critical to inform future recommendations. Although the current recommendation is to administer the vaccine in the fall or winter prior to the beginning of the RSV season, 2023-2024 vaccine administration in our cohort occurred concurrently with RSV circulation in the community, and our analysis was adjusted accordingly. It is likely that this delayed roll out after the beginning of the season reduced the total potential averted burden due to RSV vaccine last season. More so, later-vaccinated individuals received their single vaccine 10 months or more before their next likely exposure to RSV, fall of 2024. This group of vaccinees will be a valuable source of information on vaccine durability in the 2024-2025 season.

Our study is limited in its generalizability to the broader US population as evident in our higher immunization rate and relatively low rate of high-risk health conditions. Our low event rate, which is not uncommon in prospective cohort studies, precluded a fully adjusted evaluation of vaccine effectiveness and only a subset of participants had serology collected at times relevant to our analysis. This led to a smaller sample size for analysis with limited power. However, this data still provides important, preliminary observational data during the first year of vaccine implementation, and the longitudinal nature of this cohort presents the opportunity to revisit these questions in future years.

Antibody against pre-fusion RSV F is expected to have sufficient cross protection between the RSV-A and the RSV-B subgroups. To date, no major differences between the bivalent and monovalent RSV vaccine have emerged. RSV-B was the most common infecting subtype in our study, and most participants (67.5%) received the GlaxoSmithKline’s monovalent (AREXVY) vaccine, which is constructed from an RSV-A virus.^27^ Continuing genomic surveillance of RSV is key to ensuring that RSV vaccine effectiveness monitoring is robust going forward, allowing public health to respond to new strains of the virus if they emerge.

The 2023-2024 RSV vaccine program in the United States is the culmination of decades of vaccine development, and an additional vaccine has been approved as of June 2024. Over the next year, focused attention is warranted regarding the durability of vaccine-elicited immunity and the future potential for direct and indirect reduction of illness burden in all adults over 60.

## Data sharing

The data used in this study can be made available upon request. Due to Institutional Review Board (IRB) regulations, data access is controlled. Per the guidelines of the Centers for Excellence in Influenza Research and Response (CEIRR) Network, individuals seeking access must complete a data and specimen collaboration form. Requests received will be reviewed by the study investigators. For inquiries, please contact. Requests will generally receive an initial response within 3–5 business days.

## Data Availability

All data produced in the present study can be made available upon request. Due to Institutional Review Board (IRB) regulations, data access is controlled. Per the guidelines of the Centers for Excellence in Influenza Research and Response (CEIRR) Network, individuals seeking access must complete a data and specimen collaboration form. Requests received will be reviewed by the study investigators. For inquiries, please contact. Requests will generally receive an initial response within 3-5 business days.

## References

1. Havers FP, Whitaker M, Melgar M, Pham H, Chai SJ, Austin E, et al. Burden of Respiratory Syncytial Virus–Associated Hospitalizations in US Adults, October 2016 to September 2023. JAMA Network Open. 2024 Nov 13;7(11):e2444756.

2. Surie D, Yuengling KA, DeCuir J, Zhu Y, Gaglani M, Ginde AA, et al. Disease Severity of Respiratory Syncytial Virus Compared with COVID-19 and Influenza Among Hospitalized Adults Aged ≥60 Years - IVY Network, 20 U.S. States, February 2022-May 2023. MMWR Morb Mortal Wkly Rep. 2023 Oct 6;72(40):1083–8.

3. Surie D, Yuengling KA, DeCuir J, Zhu Y, Lauring AS, Gaglani M, et al. Severity of Respiratory Syncytial Virus vs COVID-19 and Influenza Among Hospitalized US Adults. JAMA Netw Open. 2024 Apr 1;7(4):e244954.

4. Van Effelterre T, Hens N, White LJ, Gravenstein S, Bastian AR, Buyukkaramikli N, et al. Modeling Respiratory Syncytial Virus Adult Vaccination in the United States With a Dynamic Transmission Model. Clin Infect Dis. 2023 Aug 14;77(3):480–9.

5. Yamin D, Jones FK, DeVincenzo JP, Gertler S, Kobiler O, Townsend JP, et al. Vaccination strategies against respiratory syncytial virus. Proc Natl Acad Sci USA. 2016 Nov 15;113(46):13239–44.

6. Melgar M, Britton A, Roper LE, Talbot HK, Long SS, Kotton CN, et al. Use of Respiratory Syncytial Virus Vaccines in Older Adults: Recommendations of the Advisory Committee on Immunization Practices - United States, 2023. MMWR Morb Mortal Wkly Rep. 2023 Jul 21;72(29):793–801.

7. Walsh EE, Pérez Marc G, Zareba AM, Falsey AR, Jiang Q, Patton M, et al. Efficacy and Safety of a Bivalent RSV Prefusion F Vaccine in Older Adults. N Engl J Med. 2023 Apr 20;388(16):1465–77.

8. Papi A, Ison MG, Langley JM, Lee DG, Leroux-Roels I, Martinon-Torres F, et al. Respiratory Syncytial Virus Prefusion F Protein Vaccine in Older Adults. N Engl J Med. 2023 Feb 16;388(7):595–608.

9. Surie D, Self WH, Zhu Y, Yuengling KA, Johnson CA, Grijalva CG, et al. RSV Vaccine Effectiveness Against Hospitalization Among US Adults 60 Years and Older. JAMA. 2024 Oct 1;332(13):1105–7.

10. Black CL, Kriss JL, Razzaghi H, Patel SA, Santibanez TA, Meghani M, et al. Influenza, Updated COVID-19, and Respiratory Syncytial Virus Vaccination Coverage Among Adults - United States, Fall 2023. MMWR Morb Mortal Wkly Rep. 2023 Dec 22;72(51):1377–82.

11. Feldstein LR, Britton A, Grant L, Wiegand R, Ruffin J, Babu TM, et al. Effectiveness of Bivalent mRNA COVID-19 Vaccines in Preventing SARS-CoV-2 Infection in Children and Adolescents Aged 5 to 17 Years. JAMA. 2024 Feb 6;331(5):408–16.

12. Monto AS, Malosh RE, Evans R, Lauring AS, Gordon A, Thompson MG, et al. Data resource profile: Household Influenza Vaccine Evaluation (HIVE) Study. Int J Epidemiol. 2019 Aug 1;48(4):1040–1040g.

13. Monto AS, Foster-Tucker JE, Callear AP, Leis AM, Godonou ET, Smith M, et al. Respiratory Viral Infections From 2015 to 2022 in the HIVE Cohort of American Households: Incidence, Illness Characteristics, and Seasonality. The Journal of Infectious Diseases. 2024 Aug 24;jiae423.

14. Petrie JG, Fligiel H, Lamerato L, Martin ET, Monto AS. Agreement between state registry, health record, and self-report of influenza vaccination. Vaccine. 2021 Sep 7;39(38):5341–5.

15. Wei J, Pouwels KB, Stoesser N, Matthews PC, Diamond I, Studley R, et al. Antibody responses and correlates of protection in the general population after two doses of the ChAdOx1 or BNT162b2 vaccines. Nat Med. 2022 May;28(5):1072–82.

16. Tenforde MW, Olson SM, Self WH, Talbot HK, Lindsell CJ, Steingrub JS, et al. Effectiveness of Pfizer-BioNTech and Moderna Vaccines Against COVID-19 Among Hospitalized Adults Aged ≥65 Years - United States, January-March 2021. MMWR Morb Mortal Wkly Rep. 2021 May 7;70(18):674–9.

17. Rolfes MA, Flannery B, Chung JR, O’Halloran A, Garg S, Belongia EA, et al. Effects of Influenza Vaccination in the United States During the 2017–2018 Influenza Season. Clinical Infectious Diseases. 2019 Nov 13;69(11):1845–53.

18. Price AM, Flannery B, Talbot HK, Grijalva CG, Wernli KJ, Phillips CH, et al. Influenza Vaccine Effectiveness Against Influenza A(H3N2)-Related Illness in the United States During the 2021–2022 Influenza Season. Clinical Infectious Diseases. 2023 Apr 15;76(8):1358–63.

19. McLean HQ, Petrie JG, Hanson KE, Meece JK, Rolfes MA, Sylvester GC, et al. Interim Estimates of 2022-23 Seasonal Influenza Vaccine Effectiveness - Wisconsin, October 2022-February 2023. MMWR Morb Mortal Wkly Rep. 2023 Feb 24;72(8):201–5.

20. Rothman KJ. Epidemiology: An Introduction. OUP USA; 2012. 281 p.

21. Bates D, Mächler M, Bolker B, Walker S. Fitting Linear Mixed-Effects Models Using **lme4**. J Stat Soft [Internet]. 2015 [cited 2024 Nov 6];67(1). Available from: http://www.jstatsoft.org/v67/i01/

22. Davison AC, Hinkley DV. Bootstrap Methods and their Application [Internet]. 1st ed. Cambridge University Press; 1997 [cited 2024 Nov 6]. Available from: https://www.cambridge.org/core/product/identifier/9780511802843/type/book

23. Canty, Angelo and Ripley, B.D. A. boot: Bootstrap R (S-Plus) Functions. 2024.

24. Havers FP, Whitaker M, Melgar M, Chatwani B, Chai SJ, Alden NB, et al. Characteristics and Outcomes Among Adults Aged ≥60 Years Hospitalized with Laboratory-Confirmed Respiratory Syncytial Virus - RSV-NET, 12 States, July 2022-June 2023. MMWR Morb Mortal Wkly Rep. 2023 Oct 6;72(40):1075–82.

25. Britton A, Roper LE, Kotton CN, Hutton DW, Fleming-Dutra KE, Godfrey M, et al. Use of Respiratory Syncytial Virus Vaccines in Adults Aged ≥60 Years: Updated Recommendations of the Advisory Committee on Immunization Practices — United States, 2024. MMWR Morb Mortal Wkly Rep. 2024 Aug 15;73(32):696–702.

26. Hall CB, Long CE, Schnabel KC. Respiratory syncytial virus infections in previously healthy working adults. Clin Infect Dis. 2001 Sep 15;33(6):792–6.

27. Ison MG, Papi A, Athan E, Feldman RG, Langley JM, Lee DG, et al. Efficacy and Safety of Respiratory Syncytial Virus (RSV) Prefusion F Protein Vaccine (RSVPreF3 OA) in Older Adults Over 2 RSV Seasons. Clin Infect Dis. 2024 Jun 14;78(6):1732–44.

