## Supplementary Appendix for "Respiratory syncytial virus (RSV) vaccine effectiveness and antibody correlates of protection among older adults in the Community Vaccine Effectiveness (CoVE) observational study"

### **Supplemental Methods**

#### **Sample selection for correlates of protection analysis**

The appropriate baseline concentration time point was selected based on the vaccination and infection status of the participant. For participants who were infected and unvaccinated, the (pre-infection) sample collected closest to the RSV infection date was selected. For those who were infected and vaccinated, the pre-infection sample was selected if the RSV vaccine was administered after the infection, while the post-vaccination sample was used if the serum specimen was collected before the RSV infection. Among uninfected participants, the pre-season sample was selected for the unvaccinated group; for vaccinated participants, the post-vaccination sample collected before or on October 25, 2023 (a week prior to the first RSV case in participants aged 60 and older), was selected.

#### **RSV preF IgG waning models**

The exponential decay mixed model after log_10_ transformation of antibody takes the following form:^1,2^

$$\log_{10}\left( RSV preF \right. \left. \mathrm{IgG}_{ij} \right)=\alpha+\beta\cdot\left( \mathrm{Time}_{ij} \right)+u_{i}+e_{ij}$$

where α and β are the fixed effects, intercept and decay rate, respectively, 𝑢*_i_* is the random intercept for each participant *i*, and 𝑒_𝑖𝑗_ are the model errors for participant *i* at time *j*. Thus, log_10_ transformation of RSV preF IgG levels is a linear function of time (i.e., steady decay rate in log scale over time).^3^

The general form of the power-law mixed model takes the following form:^1^

$$\log_{10}\left( RSV preF \right. \left. \mathrm{IgG}_{ij} \right)=\alpha+\beta\cdot\left( \log_{10}\left( \mathrm{Time}_{ij} \right) \right)+u_{i}+e_{ij}$$

where α and β are the fixed effects, intercept and decay rate, respectively, 𝑢*_i_* is the random intercept for each participant *i*, and 𝑒_𝑖𝑗_ are the model errors for participant *i* at time *j*. Thus, the log_10_ transformation of RSV preF IgG levels is a linear function of the log_10_ transformation of time (i.e., decay rates decrease over time).^3^

Natural cubic splines mixed models with two inner knots,^3^ after log_10_ transformation of antibody concentrations, were used to get the predicted curves of antibody levels for both vaccinated and unvaccinated groups. The location of the two interior knots was placed at suitably chosen quantiles of time as implemented in the splines package in R (version 4.3.0; R Core Team); the boundary knots, by default, are placed at the minimum and maximum of time. The selection of the appropriate number of inner knots (i.e., two) was done by fitting different NCS mixed models with two, three, and four inner knots, and selecting the model with *k* inner knots that best fits the data based on the Akaike Information Criterion with correction for small samples sizes (AICc) and the Bayesian Information Criterion (BIC).^4^ We then evaluated the three (ED, PL, and NCS with two inner knots) models’ fit using AICc and BIC criteria; the smaller the AICc or BIC, the better the fit. Models' predicted trajectories of RSV preF IgG levels on a log_10_ scale were plotted over time.

#### **Antibody half-life estimation**

Antibody half-life (t_1/2_), representing the number of days required to halve RSV preF IgG antibody levels, was estimated for both vaccinated and unvaccinated groups. The half-life was calculated based on the models' predictions. Specifically, for the vaccinated group, we used the baseline predicted level at 14 days post-vaccination, and for the unvaccinated group, at 1 day since the start of the study period. We then subtracted log_10_(2) to determine the predicted antibody level corresponding to a 50% reduction. By comparing the model's predicted values to the antibody level representing a 50% reduction, we identified the day (*i*-th) post-vaccination when the predicted value equaled the 50% reduction level. The half-life was then calculated by subtracting 14 days from the estimated day for the vaccinated group and 1 day for the unvaccinated group. This method was carried out for each model (ED, PL, NCS). The confidence intervals for the estimated half-lives were computed using a nonparametric bootstrap method as implemented in the boot package in R.^5,6^

#### **References**

1.	Doria-Rose N, Suthar MS, Makowski M, O’Connell S, McDermott AB, Flach B, et al. Antibody Persistence through 6 Months after the Second Dose of mRNA-1273 Vaccine for Covid-19. N Engl J Med. 2021 Jun 10;384(23):2259–61.

2.	Asamoah-Boaheng M, Grunau B, Haig S, Karim ME, Kirkham T, Lavoie PM, et al. Eleven-month SARS-CoV-2 binding antibody decay, and associated factors, among mRNA vaccinees: implications for booster vaccination. Access Microbiol. 2023 Nov 28;5(11):000678.v3.

3.	Hatzakis A, Karabinis A, Roussos S, Pantazis N, Degiannis D, Chaidaroglou A, et al. Modelling SARS-CoV-2 Binding Antibody Waning 8 Months after BNT162b2 Vaccination. Vaccines (Basel). 2022 Feb 13;10(2):285.

4.	Lüdecke D, Ben-Shachar MS, Patil I, Waggoner P, Makowski D. performance: An R Package for Assessment, Comparison and Testing of Statistical Models. Journal of Open Source Software. 2021 Apr 21;6(60):3139.

5.	Davison AC, Hinkley DV. Bootstrap Methods and their Application [Internet]. 1st ed. Cambridge University Press; 1997 [cited 2025 Mar 13]. Available from: https://www.cambridge.org/core/product/identifier/9780511802843/type/book

6.	Angelo Canty, B. D. Ripley. boot: Bootstrap R (S-Plus) Functions. 2024.
